# Spatiotemporal mapping of ex vivo myometrial contractions in adenomyosis

**DOI:** 10.64898/2026.11.10.26367720

**Authors:** Emilie Huon, Léna Zig, Olympe Séguard, Marine Plancke, Carole Abo, Marie-Charlotte Lamau, Gauthier Dietrich, Ylann Abrahami, Jordan Kouby, Julien Adam, Nicolas R. Chevalier

## Abstract

**Objective:** Uterine dyscontractility plays a major role in pain sensation, infertility, and adenomyosis pathogenesis. In vivo, contractility occurs in the inner myometrium and junctional zone, which is thickened in adenomyosis. We examined whether the junctional zone presents particular contractile features ex vivo by measuring contractility and oxytocin response in full-thickness serosa-to-endometrium myometrial slices retrieved from control and adenomyosis patients.

**Design:** Prospective ex vivo case-control study.

**Subjects:** Premenopausal women following hysterectomy: adenomyosis (n = 16) and controls (n = 12). Contractility: 57 strips (11 vs 8 patients), immunofluorescence: 10 vs 9 patients.

**Main Outcome Measures:** We resorted to spatiotemporal mapping, time-lapse video-based techniques used in the field of gastroenterology. We measured contraction frequency, amplitude, duration, velocity, site of origin, and propagation direction along the serosa-to-endometrium axis, before and after oxytocin. We measured oxytocin receptor density along the serosa-to-endometrium axis by immunofluorescence and machine learning-assisted segmentation.

**Results:** Ex vivo contractile waves propagated throughout the outer and inner myometrium, with similar amplitude and frequency. Contraction frequency, amplitude, duration, and velocity did not differ between control and adenomyosis uteri. Oxytocin increased contraction frequency and velocity and systematically caused contractile waves to originate at the inner myometrium in both groups. In both groups, oxytocin receptor density was highest in the inner myometrium. Adenomyosis patients had a 1.8-fold higher oxytocin receptor density, but a smaller oxytocin-induced increase in contraction frequency.

**Conclusion:** We provide the first ex vivo spatiotemporal time-lapse analysis of myometrial contractions. The whole myometrium contracts ex vivo, not just the inner part (junctional zone). This uniform myogenic motility is presumably inhibited neurally in vivo in the outer myometrium. Ex vivo adenomyosis basal contractility was similar to that of controls. Adenomyotic myometrium overexpressed oxytocin receptors compared to controls, but this did not translate into a stronger contractile response to oxytocin. In both groups, oxytocin caused contractile waves to originate at and propagate from the sub-endometrial myometrium, where receptor density is higher. This demonstrates that contractile wave direction can be controlled hormonally.

## Introduction

Uterine contractions depend on hormonal, mechanical, and inflammatory signals (1–3). In severe dysmenorrhea, endometriosis, and adenomyosis, this contractility becomes disrupted, leading to hypercontractility and dysperistalsis (4–8). In healthy eumenorrheic women, uterine contractions (peristaltic waves) concentrate in the inner myometrium(9–11), originating in the junctional zone (JZ) (12–14). The characteristic low T2 of the JZ remains visible on MRI of excised uteri, although less sharply than in vivo (15,16). Uterine peristaltic contractions vary in direction, frequency, amplitude, and velocity depending on cycle phase to facilitate sperm transport (cervix-to-fundus direction during ovulation) or menstrual fluid evacuation (fundus-to-cervix direction during menses) (6,17,18).

Endometriosis is associated with a significantly increased peristaltic activity (5,19,20). This excessive peristalsis generates elevated uterine pressures that can promote retrograde transport of endometrial fragments through the fallopian tubes into the peritoneal cavity (21). This mechanism is considered a crucial early step in disease pathogenesis (6,21). Adenomyosis is characterized by the presence of endometrial glands within the myometrium, and by a thickened (>12 mm) JZ (12,22,23). Contractions in adenomyosis are uncoordinated, which may disrupt sperm transport and contribute to infertility (24–26). A study on isolated myometrial strips found that adenomyotic tissue exhibits markedly higher spontaneous contraction amplitude compared to healthy tissue (27). Chronic dyscontractility could induce JZ microtrauma that favours endometrial invasion of the myometrium, a hypothesis known as “tissue injury and repair” (28). The underlying causes of contractile dysfunction in adenomyosis remain poorly understood.

In vivo cine MRI / TVUS captures contraction direction, velocity, and coordination, but does not have access to force or pressure. Conversely, ex vivo strip studies report force but obliterate wave-like aspects of contraction propagation. In vivo, uterine contractility is found to be restricted to the inner myometrium, which encompasses the JZ (5,9–11). It is not known whether the JZ or the inner myometrium exhibit distinct contractile features ex vivo, in an organ bath. To address these questions and methodological gaps, we developed here an optical, ex vivo approach to measure contractile wave origin, amplitude, velocity, direction, and frequency across the entire myometrial thickness, from serosa to endometrium. Our method draws on tissue displacement mapping techniques previously applied to intestinal smooth muscle (29,30), and on speckle-tracking methods used in in vivo uterine studies (31). Because oxytocin receptors (OTR) have previously been found to be overexpressed in adenomyosis myometrium (27,32), we also tested the contractile response to the uterotonic hormone oxytocin (33,34), and histologically characterized the OTR distribution across the myometrium of adenomyosis and control uteri.

## Materials and Methods

### Patients and tissue collection

Myometrial biopsies were obtained after hysterectomy from 28 premenopausal women at Hôpital Paris Saint Joseph. Patient characteristics are detailed in Table 1. Ethical approval for the study was granted by the Groupe Ethique Recherche Médicale (GERM) under approval number IRB 00012157. All patients were provided with written information about the study and consented to participate.

**Table 1:** Patient cohort description. Clinical characteristics of patients who underwent hysterectomy in the ex vivo contractility cohort (n = 19: 11 adenomyosis, 8 controls) and the immunofluorescence cohort (n = 19: 10 adenomyosis, 9 controls). Quantitative variables are expressed as median [interquartile range] and qualitative variables as % (N). Secondary pathology refers to a condition diagnosed alongside the primary indication for hysterectomy. Groups were compared with Welch’s t-test (age) and Fisher’s exact test (parity, hormonal treatment). Cycle phase and pathologies are reported descriptively.

|  | Ex vivo |  |  | Immunofluorescence |  |  |
|---|---|---|---|---|---|---|
| Variables | Case<br>n = 11 | Controls<br>n = 8 | p value | Case<br>n = 10 | Controls<br>n = 9 | p value |
| Age (median) [IQR] | 49 [44-53] | 45.5 [43.3-49.8] | 0.28 | 45.5 [41-53.3] | 47 [41.5-50] | 0.69 |
| Parity % [N] |  |  | > 0.99 |  |  | > 0.99 |
| — Nulliparous | 18.2 [2] | 12.5 [1] |  | 20 [2] | 11.1 [1] |  |
| — Parous (≥1) | 72.7 [8] | 87.5 [7] |  | 80 [8] | 77.8 [7] |  |
| Hormonal treatment % [N] | 36.3 [4] | 25 [5] | 0.37 | 30 [3] | 22.2 [2] | > 0.99 |
| Menstrual cycle % [N] |  |  | — |  |  | – |
| — Unknown | 18.2 [2] | 52 [16] |  | 60 [6] | 11.1 [1] |  |
| — Menses | 9.1 [1] | 15 [3] |  | 0 [0] | 22.2 [2] |  |
| — Proliferative phase | 18.2 [2] | 15 [3] |  | 10 [1] | 22.2 [2] |  |
| — Secretory phase | 18.2 [2] | 15 [3] |  | 10 [1] | 22.2 [2] |  |
| Pathology % [N] |  |  |  |  |  |  |
| — Adenomyosis | 100 [11] |  |  | 100 [10] |  |  |
| — Fibroids |  | 100 [8] |  |  | 100 [9] |  |
| Secondary pathology % [N] |  |  |  |  |  |  |
| — None | 27.3 [3] | 87.5 [7] |  | 30 [3] | 88.9 [8] |  |
| —Endometriosis | 0 [0] | 0 [0] |  | 30 [3] | 0 [0] |  |
| — Fibroids | 63.6 [7] |  |  | 30 [3] |  |  |
| — Hyperplasia | 9.1 [1] | 12.5 [1] |  | 10 [1] | 0 [0] |  |
| — Adenoma | 0 [0] | 0 [0] |  | 0 [0] | 11.1 [1] |  |

Two adjacent full-thickness biopsies (∼30 × 10 × 10 mm, serosa to endometrium) were cut out from the anterior wall, midway between cervix and fundus, avoiding any visible myomas. The first biopsy was immediately fixed in 4% PFA overnight for immunofluorescence (n = 10 adenomyosis, n = 9 controls). The second biopsy was transported in ice-cold medium (DMEM:F-12 + GlutaMAX, Gibco, Thermo Fisher) pre-oxygenated with carbogen (95% O2, 5% CO2). Ex vivo contractility assays were performed on this second biopsy, within 6 hours of retrieval (n = 11 adenomyosis, n = 8 controls). Ten patients (5 per group) contributed to both ex vivo contractility and immunofluorescence analysis.

### Ex vivo contractility

Each fresh biopsy was further sliced into three ∼1 mm thick strips (final size ∼30 × 10 × 1 mm), with circumferential fibers running along the strip width. Carbon black microparticles were adhered to each slice to serve as fiducial markers, and the 3 strips were pinned vertically (serosa at the top, endometrium at the bottom), next to each other, in a Sylgard-lined container at 37°C, continuously bubbled with carbogen (Figure 1A,C, Video S1). Once spontaneous phasic contractions appeared (after 1-3 h), they were recorded at 1 frame per second (1Hz) with a Basler acA3800 camera mounted with a 25mm lens. Recording times were 20 min for baseline and 30 min after 10 µM oxytocin (Sigma O4375) administration. For each patient, contractile behaviour was averaged over all contractions in the recording intervals and over the three strips, thus reducing intra-patient technical variability.

**Figure 1.**
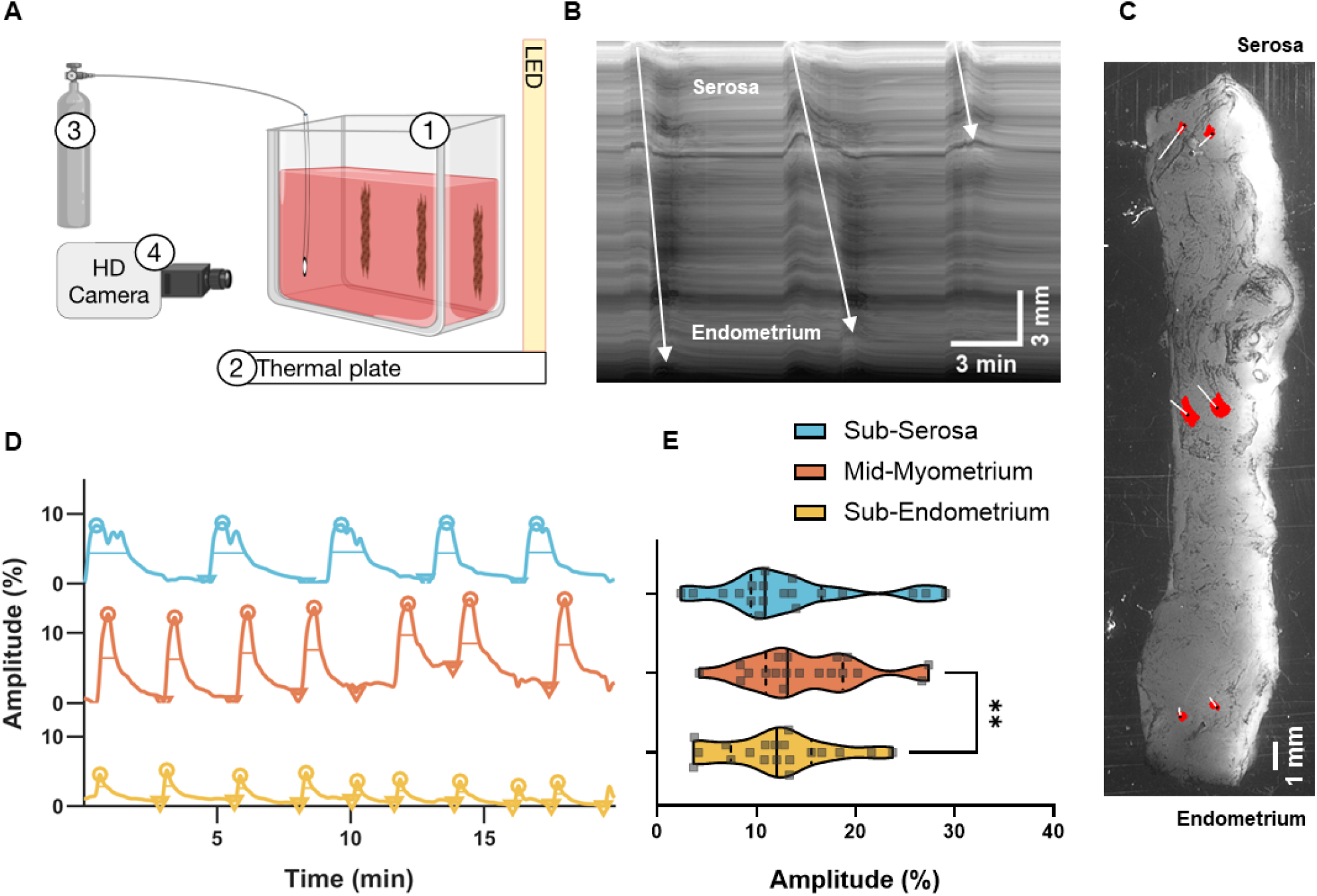
Uterine contractile waves propagate with uniform amplitude across the whole myometrial thickness. (A) Experimental setup. Three full-thickness strips per patient are pinned vertically in an organ bath (1), thermoregulated at 37°C (2), continuously bubbled with carbogen (3), and recorded with a video camera (4) at 1 Hz for up to an hour. (B) Representative 20-min kymograph showing three serosa-to-endometrium waves (arrows). (C) A strip covered with carbon black particles for displacement tracking (still shot from Video S1). Shown in red are the successive positions of three pairs of points (at the sub-serosa, mid-myometrium, and sub-endometrium), tracked over 20 min. (D) Representative deformation traces of the three pairs of points at the sub-serosa (blue), mid-myometrium (orange), and sub-endometrium (yellow). Frequency was derived from detected peaks (○), amplitude as peak-to-preceding-valley (▽) difference, and duration as width at half-prominence (-). (E) Contraction amplitude at different positions along the myometrium, for all patients (n = 19: 11 adenomyosis, 8 controls). Each point is the mean of three slices per patient (∼5 contractions per slice per recording). Repeated-measures one-way ANOVA with Tukey’s post-hoc test; **p = 0.0048.

### Video analysis

We tracked displacement of carbon black particles adhered to the myometrium with the TrackV2 ImageJ plugin. Three pairs of particles were selected for each strip, at the sub-serosal, mid-myometrial, and sub-endometrial levels (Figure 1C). For each pair, we computed the deformation relative to the relaxed state: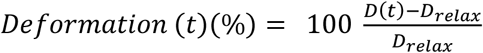, where D(t) is the inter-point distance (along x, i.e. perpendicular to the serosa-endometrium axis), and *D*_*relax*_the maximum interpoint-distance reached during the recording. Traces were smoothed with a Gaussian filter (σ = 3 s), and contractions were detected using custom MATLAB peak-detection algorithms (https://github.com/emiliehuon/uterus_analysis_contractility), adjusting the minimum peak prominence for each recording (0.1 – 1 %) to exclude noise. We computed the frequency (peaks per 10 min), amplitude (peak-to-preceding valley, %), and duration (width at half-prominence, min).

For each propagating wave, its site of origin was measured directly from the video and normalised to strip length. Velocity was measured as the distance travelled by the contractile wave divided by the travel time. Sites of origin and velocity were averaged over all visible waves for each strip, and then over the three strips for each patient. Other contraction parameters were averaged over the three pairs of points, and over the three strips (i.e., 9 measurements per patient).

### Immunofluorescence

The fixed biopsy was cryoprotected in 30% sucrose, embedded in PolyFreeze (Sigma), and cut in 18 µm sections deposited on Superfrost glass slides. Slides were permeabilized and blocked in PBS, 1% BSA, and 0.1% Triton for 1 h, incubated overnight with Oxytocin receptor antibody (OTR, LS-A246, LSBio, 1:100), thoroughly washed, and further incubated in Alexa Fluor 647 anti-rabbit IgG (Thermo Fisher, 1:400, 3 h). Slides were kept under gentle agitation throughout to optimize staining homogeneity. They were finally washed, cover-slipped, and imaged at 10x on a Zeiss wide-field epifluorescence microscope (ImagoSeine platform), in mosaic mode to capture the full fluorescence distribution from serosa to endometrium. Mosaic tiles (∼100-200 per sample) were stitched with ZEN software (Zeiss).

### Image Analysis

Ten ROIs (each 780 µm x 780 µm) were placed along the serosa-endometrium axis, avoiding large vessels, holes, and folds, using a custom Fiji macro (GitHub, as above). OTR-positive pixels were segmented with Trainable Weka Segmentation(35). OTR density was expressed as the OTR-positive area divided by the ROI area. ROI positions along the serosa-to-endometrium axis were normalised, and a smooth expression profile fitted through the 10 points. The average OTR density was computed as the area under the smoothed expression profile.

### Statistical analysis

Statistical analyses were performed under GraphPad Prism 11. Tests are specified in figure and table legends. Data are reported as median [IQR], or % (N); p < 0.05 was considered significant.

## Results

### Characteristics of ex vivo spontaneous myometrial contractile waves

We characterized the contractile behaviour of full-thickness – from serosa to endometrium - human myometrial strips (Figure 1). Contractile activity occurred mostly as propagating contractile waves that travelled along the slice length, from the serosa to the endometrium, or in the opposite direction (Figure 1B-D, Video S1). The cross-sample average (n = 19) contraction frequency was 2.3 cycles per 10 minutes (cp10m), amplitude 13 %, duration 1.0 min, and velocity 1.4 cm/min. Importantly, contraction frequency and amplitude were homogeneous along the slice length (Figure 1D, E): the outer myometrium contracted as much as the inner myometrium in our ex vivo organ bath.

Individual contractile waves originated at various positions along the slice length. Waves that originated at the serosa travelled predominantly towards the endometrium, and conversely those nucleating at the endometrium propagated to the serosa; waves originating in the middle of the preparation travelled in both directions. This behaviour was similar to that of myogenic contraction waves in the intestine (36). The cross-sample average normalized origin of contraction was 0.52 (0 = endometrium | 1= serosa; n = 19), i.e., close to the mid-myometrium.

Spontaneous contraction frequency, amplitude, duration, and velocity did not differ between control and adenomyosis uteri (Figure 2A-D). Contractile waves tended to originate closer to the endometrium in adenomyosis than in controls (Figure 2F), but the difference was not statistically significant.

**Figure 2.**
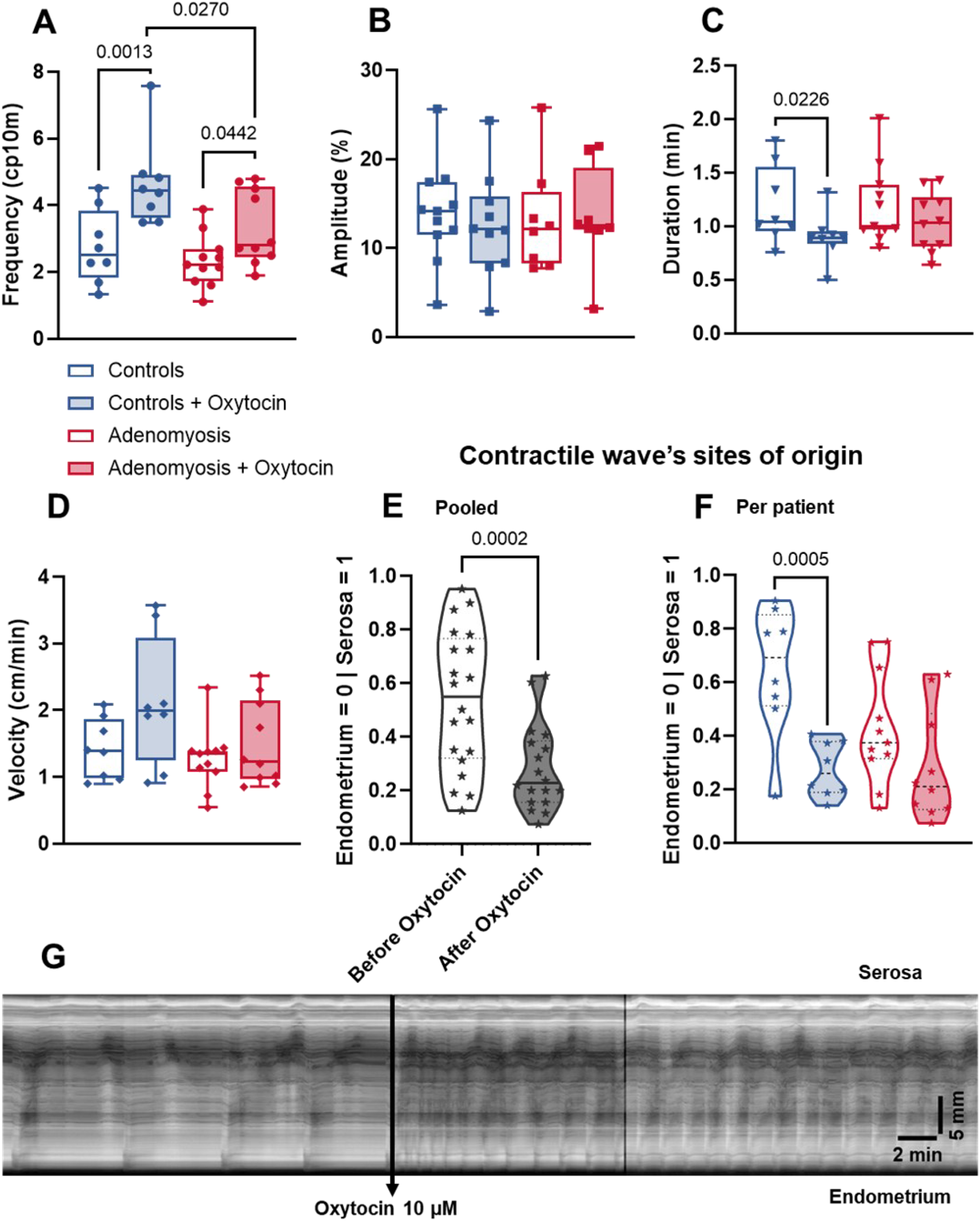
Effects of oxytocin on the contractility of control and adenomyosis myometrium. (A-F) Contractile parameters before and 7-30 min after oxytocin, in controls (blue, n = 8) and adenomyosis (red, n = 11); open boxes before oxytocin, filled boxes after oxytocin. Each point represents one patient (mean of 5.0 ±1.8 contractions per strip, and for three strips per patient). Two-way ANOVA (group x period, repeated measures on period), mixed-effects model, followed by Sidak’s post-hoc test. Comparisons not indicated were not significant. (E, F) The sites of origin of the contractile waves are shown for all patients, with adenomyosis patients separated from controls. (G) Representative kymograph of a myometrial strip along the serosa-to-endometrium axis, showing contraction frequency and direction change after administration of 10 µM oxytocin.

### Oxytocin-induced contractility in adenomyosis and control patients

Because oxytocin receptors have been previously reported to be overexpressed in adenomyosis myometrium (27,32), we next examined the contractile response of control and adenomyosis tissue to 10 µM oxytocin. This concentration was chosen because it elicited pronounced changes in motility, which were difficult to detect at lower concentrations (≤1 µM); non-pregnant hysterectomy myometrium is known to be much less oxytocin-sensitive than pregnant cesarean section myometrium (33,37,38). Oxytocin induced time-dependent alterations. Right after administration, it caused a transient (0-7 min) state of “tremor”, characterized by short-duration, low-amplitude, high-frequency contractions (Figure 2G, Video S1, Figure S1). At longer times after administration (7-30 min), the slice reached a stationary state where contractions were more frequent than before administration (Figure 2A), but less frequent than in the transient “tremor” episode (Figure S1). While oxytocin did not affect wave amplitude (Figure 2B, Figure S1), it increased wave velocity (Figure 2D, Figure S1) and concomitantly decreased contraction duration (Figure 2C, Figure S1). Changes induced by oxytocin were similar in control and adenomyosis patients (Figures 2B-E) except for frequency, where control uteri exhibited a stronger response than adenomyosis (Figure 2A).

A striking effect of oxytocin was that it consistently shifted the site of origin of contractile waves towards the endometrium (Figure 2E). This shift was present in both patient groups, but was more pronounced in controls (Figure 2F). This resulted in a majority of waves traveling from the endometrium towards the serosa after oxytocin administration (Figure 2G, Video S1).

To understand the molecular origin of this change, we mapped the density of OTR receptors along the serosa-to-endometrium axis (Figure 3A-D). We found that OTR expression continuously increased from the serosa to the endometrium (Figure 3A,E): OTR expression was highest in the sub-endometrial myometrium in both patient groups. Adenomyosis patients showed a significantly higher average OTR density, 1.8-fold, than controls (Figure 3F).

**Figure 3:**
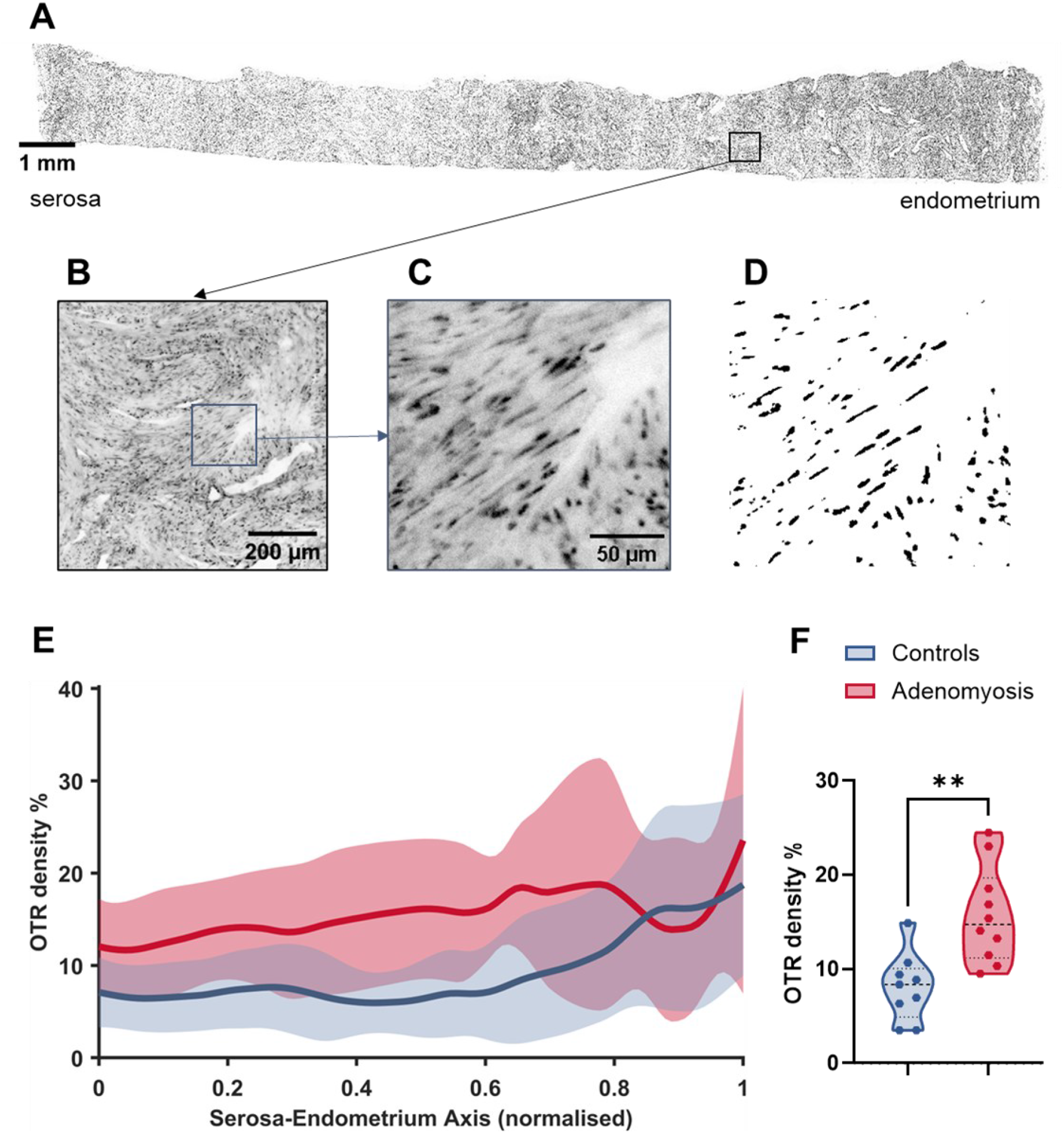
Oxytocin receptor density follows an increasing serosa-to-endometrium gradient and is higher in adenomyosis patients. (A) Full-thickness myometrium section immunostained for OTR, mosaic of ∼200 stitched tiles. (B) Magnified view of one ROI used for analysis (780 x 780 µm; black square in A). (C) Magnified view of the raw signal (blue square in B). (D) Binary obtained by thresholding the Trainable Weka Segmentation probability map in Fiji. (E) OTR density profiles ± SD along the serosa-to-endometrium axis (0 = serosa | 1 = endometrium). (F) Integrated OTR density (area under each profile) for adenomyosis (n = 10) and controls (n = 9). Violin plots with median and quartiles; Welch’s t-test, **p = 0.0015.

## Discussion

Most uterine contractility organ bath studies focus on the force signal, without looking at the strip itself. In this study, we harnessed optical tracking techniques commonly used in the assessment of intestinal motility to monitor the contractility of full-thickness myometrium strips. This revealed key *spatiotemporal* - as opposed to just temporal - information about myometrial contractility. This approach allowed us to compare outer vs inner myometrium and to derive data pertaining to contractile wave initiation and propagation. The baseline ex vivo frequencies we find (∼2 cycles / 10 min) are similar to ex vivo values reported with force transducers (33,39), but are ∼5-10 times lower than in vivo values (10,17,18).

### The whole myometrium contracts ex vivo, not just the JZ

MRI (4) and ultrasound (9–11,40) investigations have consistently shown that, in vivo, only the inner myometrium contracts (is strained, deformed). In contrast, in our ex vivo study, we found that outer and inner myometrium displayed similar contractile amplitude and frequency (Figure 1): contractile waves propagated seamlessly from one region to the other, and could alternatively originate in the outer or inner myometrium (Figure 2, Video S1). How do we reconcile these apparently contradictory in vivo and ex vivo observations? We have previously shown that ex vivo uterine contractility is insensitive to tetrodotoxin (41), i.e., it is *myogenic*. Although nerve fibers are still present in the tissue we monitor, they have been severed from their nerve cell bodies in the hypogastric plexus (42) during the hysterectomy operation and are *de facto* no longer functional. In a forthcoming publication, we show that adrenergic inhibitory innervation is mostly concentrated in the outer myometrium: it is reasonable to conclude that this innervation actively quenches, in vivo, the potential myogenic contractility of the outer myometrium.

### Oxytocin receptor distribution can control contractile wave direction

At baseline, contractile waves originated at various sites along the myometrium thickness, either in the outer or the inner myometrium. After oxytocin administration, contractile waves systematically originated in the inner myometrium (close to the endometrium) in both patient groups (Figure 2), and thereafter propagated to the outer myometrium. We further found that this inner myometrium region corresponds to the one where OTR expression is highest (Figure 3). This suggests that the spatial distribution of OTR can determine the preferential site of origin of contractions and their ensuing direction. It is known that oxytocin injection modulates the frequency and direction of uterine peristalsis in vivo (6,18), and that OTR expression fluctuates across the menstrual cycle (43). A differential expression of OTR at the fundus vs cervix at different phases of the menstrual cycle could serve as a guiding cue for cervico-fundal vs fundo-cervical waves (32,44).

### Adenomyosis vs control: more oxytocin receptors but similar myogenic contractility

We found that spontaneous contraction frequency, amplitude, duration, and velocity did not differ between adenomyosis patients and controls. This is consistent with a recent in vivo ultrasound study of adenomyotic patients (24) that found only minor alterations to frequency, amplitude, duration, and velocity, but a significant alteration of contraction coordination. In contrast, Guo et al. (27) reported higher spontaneous amplitude in ex vivo strips from adenomyosis patients. Their measurements were restricted to the outer myometrium of patients with highly symptomatic adenomyosis, whereas we averaged contractility over the whole myometrial thickness in a cohort that was not selected based on symptom severity. Our results indicate that the spontaneous *myogenic* contractility of adenomyosis myometrium does not significantly differ from that of controls: our experiment does not capture alterations of the *neurogenic* component, which are, as we will show in a forthcoming publication, substantial.

We found that OTR density was ∼2 times higher in adenomyosis than in controls (Figure 3), in agreement with previous reports of OTR overexpression in adenomyotic myometrium (27,32). Although we did not assess OTR distribution along the cervico-fundal axis, it is possible that further alterations in the spatial distribution of these receptors could lead to a loss of contractile wave coordination and direction (24,32,45).

Administration of 10 µM oxytocin increased contraction frequency in both patient groups. The frequency increase was, however, not correlated with OTR expression: it was 1.6-fold lower in adenomyosis uteri than in controls, whereas OTR density was 1.8 times-fold higher in adenomyosis patients than in controls. It is likely that the concentration we used (10 µM) is saturating, i.e., it yielded a response independent of OTR expression. Dose-response experiments could reveal a sensitivity to lower oxytocin concentrations in adenomyosis uteri. It is also possible that receptor overexpression does not translate to a stronger response if receptors are desensitised or internalised, or if downstream signalling is altered (33,34).

### Study limitations

Several limitations of this study should be acknowledged. First, our sample size was modest (n = 8-11 per group), which limited statistical power: smaller between-group differences in oxytocin-induced contractility may have gone undetected. Many novel results were, however, common to both groups and therefore based on a more substantial number of patients (n = 19 total). Second, we extracted contractile signals from operator-defined pairs of points: a more sophisticated approach would consider the full deformation field of the slice (31). Third, we used a single oxytocin concentration and did not obtain full dose-response curves for adenomyosis vs control tissue. These data will be provided in a future report. The high concentration used (10 µM) appeared necessary to elicit changes in contractility in the non-pregnant, hysterectomy myometrium we examined. The literature confirms that non-pregnant myometrium is much less sensitive to oxytocin than pregnant caesarean-section tissue (33,37,38). Fourth, most of the adenomyosis tissue we collected did not present with adenomyosis lesions in the particular biopsy we retrieved; our data therefore pertains to potential systemic (myometrium-wide) alterations to myogenic contractility, rather than local alterations induced specifically by lesions.

## Conclusion

We provided spatially resolved measurements of contractility across the whole thickness of the non-pregnant human myometrium, from serosa to endometrium. Ex vivo contractility is purely myogenic, and myogenic motility was similar in the outer, middle, and inner myometrium. This sharply contrasts with the in vivo (MRI, TVUS) observation that only the inner myometrium is involved in contractile activity. We suggest that the difference resides in the *neuro-myogenic* activity of the uterus in vivo, i.e., active quenching of contractions by inhibitory nerve activity. Oxytocin (10 µM) caused contractile waves to originate in the inner myometrium close to the endometrium, in a region where OTR density was highest. This indicates that OTR spatial heterogeneity in the uterus can control the direction of uterine contractile waves during the menstrual cycle. Although adenomyosis uteri exhibited higher OTR expression, their spontaneous and oxytocin-induced myogenic contractile behaviour did not differ substantially from that of control samples.

## Data Availability

All data produced in the present study are available upon reasonable request to the authors

https://github.com/emiliehuon/uterus_analysis_contractility),

## Acknowledgement

We thank Olivier Cardoso (MSC laboratory) for developing and sharing the TrackV2 plugin. We thank Thomas Rios, Marine Panza and Guillaume Quibeuf for their help with oxytocin receptor imaging and acknowledge the ImagoSeine core facility of the Institut Jacques Monod, member of the France BioImaging infrastructure (https://ror.org/01y7vt929) supported by the French National Research Agency (ANR-24-INBS-0005 FBI BIOGEN) and GIS-IBiSA. We thank the anatomopathology staff of Hôpital Paris Saint-Joseph: A. Gérard, G. Toutain, M. Peronnet and A.Bibollet, Olivia and S. Dequeant.

