## Supplemental Figure S1 for "Spatiotemporal mapping of ex vivo myometrial contractions in adenomyosis"

### Supplementary materials

#### 1. Video S1

**Video S1 – Time-lapse recording (50 min) of a myometrial strip** from a patient with adenomyosis, 20 min before and 30 min after addition of 10 µM oxytocin (time 0s). Serosa at the top, endometrium at the bottom. Acquired at 1 frame/s and played at 60 frame/s (60x real time). The kymograph of the strip's left side is shown in Figure 2G.

50 min recording of one slice from an adenomyosis patient. Kymograph of the same video (left side) is shown in Figure 2G. 20 min before oxytocin addition and 30 minutes after addition (including short- and long-term). Serosa on top, endometrium at bottom. 60 frames per second.

#### 2. Figure S1

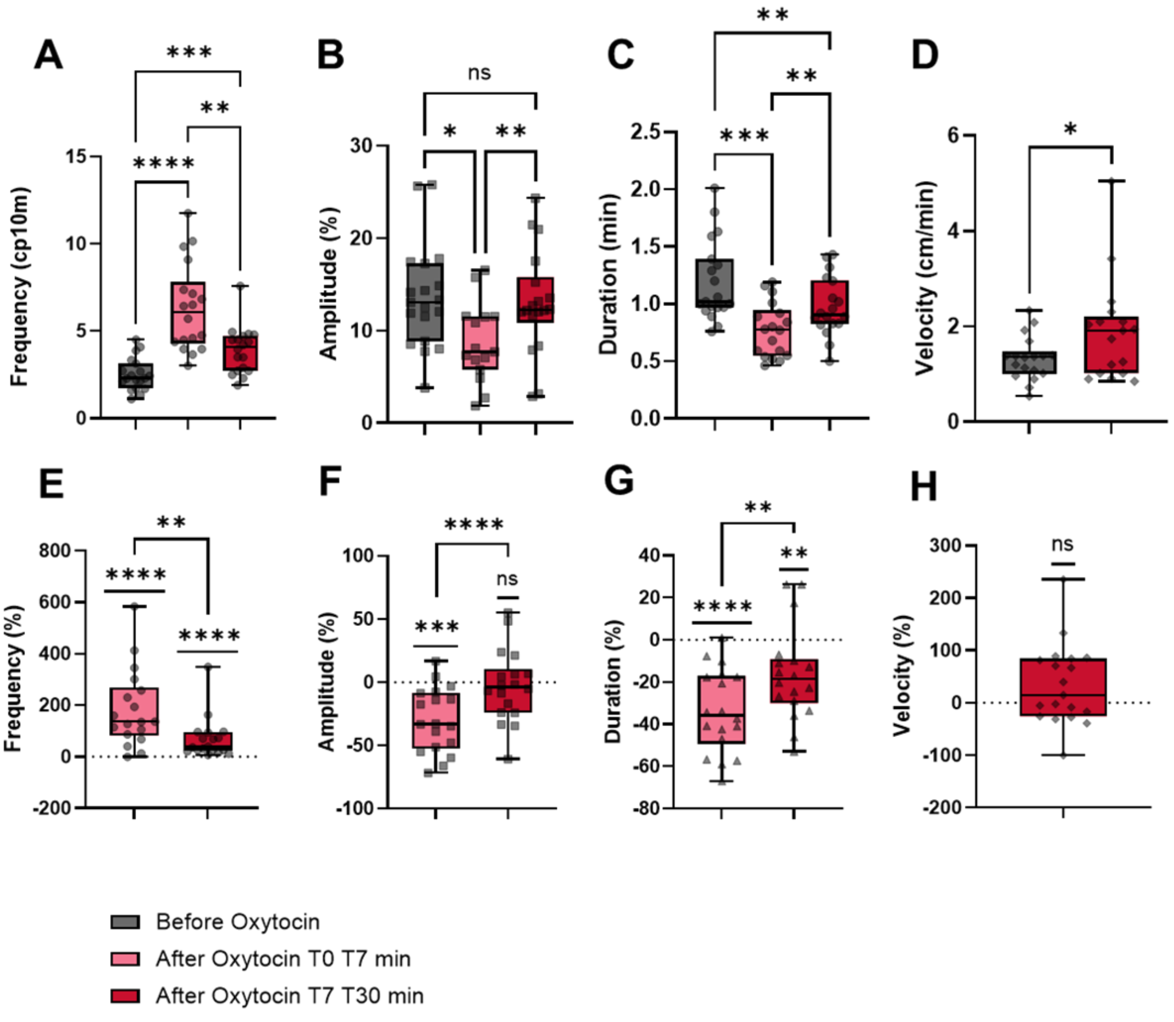


**Figure S1 – Short- and long-term effects of oxytocin on myometrial contractility, all patients pooled (adenomyosis and controls, n = 19).** Each point represents one patient (mean of 5.0 ±1.8 contractions per strip, and for three strips per patient). **(A-C)** Contraction frequency, amplitude, and duration before, 0-7 min, and 7-30 min after 10 µM oxytocin. Repeated-measures mixed-effects one-way ANOVA, followed by Tukey's post-hoc test. **(D)** Wave velocity (cm/min) before and 7-30 min after oxytocin, paired t-test. **(E-H)** Same parameters expressed as percentage change from baseline. Each period was compared with zero using a one-sample t-test, and the two periods were compared using the Wilcoxon matched-pairs signed-rank test. Comparisons not indicated were not significant; $p < 0.05$ was considered significant.
